# Execution by lethal injection: Autopsy findings of pulmonary edema

**DOI:** 10.1101/2022.08.24.22279183

**Authors:** Joel B. Zivot, Mark A. Edgar, David A. Lubarsky

**Author notes:** (Corresponding Author) Joel B. Zivot, MD, FRCP(C), MA. Authors’ contributions: All authors contributed equally to the creation of this manuscript. Role of funding source: No funding source to disclose. Ethics committee approval: N/A (excluded from review according to applicable HIPPA rules).

## Abstract

Lethal injection is the common method of capital punishment in the United States (US), but problematic executions feed controversy centered on the 8^th^ amendment of the US constitution that proscribes cruel punishment.^1^ However, the definition of cruelty is contentious, and lethal injection displays few signs of the hidden course of a potentially cruel death. Few survive and those who die cannot give witness. In this study, we reviewed autopsy reports of prisoners executed by intravenous midazolam (usually followed by a neuromuscular blocking agent and potassium chloride) or pentobarbital as a single drug. Autopsy reports for 43 inmates obtained from eight states were reviewed to identify any abnormal findings. Pulmonary edema (often fulminant with froth filling airways) emerged as a consistent and unexpected abnormality in 33/43 autopsies (76.74%), 23/28 midazolam executions (82.14%), and 10/15 (66.66%) pentobarbital executions. Injecting heavy overdoses of either acidic or basic solutions into the bloodstream may be directly toxic to pulmonary capillary endothelial cells. The injury manifests by the sudden escape of edema fluid into normally air-filled lungs, resulting in pulmonary edema. These findings explain witnessed respiratory distress during executions and increase concern about an inability of an execution witness to observe such a finding when a paralytic is added.

## Introduction

In the United States (US), lethal injection is currently the most common method of state-sponsored judicial execution. Forensic pathologist, Dr. Jay Chapman, originally proposed a three-drug protocol^2^ involving a drug to render the inmate unconscious followed by a neuromuscular blocking agent to paralyze the muscles of respiration and potassium chloride to stop electrical activity in the heart as a more humane alternative to death in the electric chair. Lethal injection protocols have varied over time in the US, but a three-drug protocol initiated with a large intravenous dose midazolam (usually 500 mg) has been used in seven states and a protocol employing pentobarbital (usually as a single agent in a dose of 2500 mg or 5000 mg) has been employed in 14 states. Protocols involving midazolam have generated controversy with eyewitness reports of movement following administration of the drug and observations indicating respiratory distress (grunting, straining, stridor, gurgling, coughing).

Lethal injection, as a lawful form of capital punishment in the United States, meets its purpose when the method of execution succeeds in killing the inmate. While the pharmacokinetic and pharmacodynamics of the drugs have been either directly studied or postulated^3 4^, structural effects of drugs employed in the specific context of lethal injection have not been investigated until now. Death certificates provide information on what contributes to the cause of death but in lethal injection, the cause of death is always listed as ‘homicide’^5^, a legal term for a human death caused by a human, and can be legal or not. The purpose of this study was to review autopsy reports of 43 inmates executed by lethal injection using two separate protocols, one that includes midazolam and the other utilizing pentobarbital as the single agent, in order to investigate the anatomic effects of execution by lethal injection.

## Materials and Methods

Autopsy reports were obtained from eight states (Alabama, Arkansas, Arizona, Florida, Georgia, Ohio, Oklahoma, and Virginia) for inmates executed by lethal injection protocols that included either pentobarbital alone (15 cases) or midazolam accompanied by one or more additional drugs that include Potassium Chloride/Acetate and a paralytic such as Vecuronium Bromide (28 cases). One of the autopsies (case 24) was performed in Ohio by one of the authors (ME). Autopsies are in the public domain and are not considered part of PHI and are therefore not constrained for release by HIPPA. The final data set under review in this series does not include the names of any individuals. Midazolam was either administered as the first of three drugs (followed next by a neuromuscular blocking agent such as vecuronium bromide and then potassium chloride/acetate in 27 cases) or in combination with hydromorphone (one case). Gross descriptions of organs described in each report were reviewed for evidence of any abnormality. Microscopic descriptions (available in 11 midazolam executions) were also reviewed. Detailed timelines released by the State were available for 22 midazolam executions, some of which were accompanied by witness observations made at the time of execution by attorneys or representatives of the media. Postmortem interval was calculated using time of death (either obtained from state records or media reports) and time of autopsy.

## Results

Inmates ranged in age from 35 to 83 years in age; 42 were male and 1 female. Where known, postmortem intervals ranged from 4 hours and 22 minutes to 2 days; postmortem intervals were not known for pentobarbital executions. The autopsy was conducted the day following execution in 4 cases, but postmortem interval could not be calculated from available data.

Gross descriptions varied in level of detail. For example, cardiac valve circumferences were measured in some cases and not others. There were most often two intravenous lines in place (one in each antecubital fossa), but additional puncture wounds and/or vascular access catheters were described in some cases, with a maximum of five intravenous catheters in place (right and left antecubital, right hand, left wrist and right groin).

Among the expected findings for this population were varying degrees of coronary atherosclerosis (with up to 75% stenosis of left anterior descending coronary artery), hepatomegaly, and an incidental thyroid mass, but no consistent changes were found in organ systems other than respiratory. A cerebellar tumor was noted in case 10, but histologic findings were not reported. Blood was described as “present from the mouth” in one midazolam execution, and in three others there was either watery or bloody fluid in the mouth and nose. An unexpected macroscopic finding in 33 of 43 autopsies (24 midazolam and 10 pentobarbital executions) was the presence of fulminant pulmonary edema as evidenced by the presence of froth, frothy fluid or blood-tinged froth located in the in the tracheobronchial tree (8 midazolam and 6 pentobarbital executions) or fluid in lung parenchyma or small airways (15 midazolam and 4 pentobarbital executions). The presence of pulmonary edema was confirmed microscopically in 8 midazolam autopsies, including one case in which there was no macroscopic evidence of pulmonary edema. Altogether, 34 of 43 cases showed some evidence of edema in lung parenchyma or airways. Lung weights ranged from 430 to 1150 grams for the right (mean 765 grams) and from 460 to 980 grams for the left (mean 681 grams).

Microscopic examination of brain tissue was performed in 4 cases; 3 cases (cases 8, 22, and 24) reported histologic evidence of hypoxic-ischemic neuronal injury and one showed no histologic abnormality in the brain. (See table 1)

**Table 1:** Pathologic findings and autopsy and pertinent witness observations for 43 inmates executed with lethal injection protocols using midazolam. Abbreviations: HIE: hypoxic-ischemic encephalopathy; HM: hydromorphone; KCL: potassium chloride; Micro: microscopic findings; MZ: midazolam; N/A: not available; PB: pentobarbital; PMI: postmortem interval; RBC: red blood cells.

| Inmate | State | R lung | L lung | Micro | Drug(s) | Observations | PMI |
| --- | --- | --- | --- | --- | --- | --- | --- |
| 1 | AL | 598 g; parenchyma oozing moderate amounts of yellow-tinged frothy fluid | 470 g | None noted | 3 drug MZ | Shallow breathing for 10 min after administration of MZ | 7 h 45 min |
| 2 | FL | 555 g; congestion and edema | 680 g | None noted | 3 drug MZ | None available | 10 h 45 min |
| 3 | AL | 1020g; major bronchi contain froth; lung oozing large amounts of yellow-tinged frothy fluid | 870 g | None noted | 3 drug MZ | Rapid breathing; chest heaved up and down (for at least 3 minutes) | 13 h 22 min |
| 4 | FL | 900g; frothy red fluid oozes from surface on manual compression | 880 g | Acute pulmonary congestion and edema | 3 drug MZ | Feet twitched, yawned | 14 h 58 min |
| 5 | FL | 610 g; nonspecific congestion | 565 g | None noted | 3 drug MZ | Body convulsed for 10 seconds; eyes fluttered; mouth opened | 13 h 24 min |
| 6 | FL | 1120 g; ooze froth with slight manual compression; marked congestion and edema | 965 g | None noted | 3 drug MZ | Chest heaved up and down for more than 5 minutes; at one point breath grew shallow | 14 h 17 min |
| 7 | AL | 1150 g; upper airways contain blood-tinged frothy edema fluid; intra-pulmonary airways show blood-tinged edema fluid | 910 g | None noted | 3 drug MZ | Heavy breathing | 12 h 32 min |
| 8 | VA | 886 g; upper airways foamy liquid | 715 g | Lung: edema and RBC; brain: HIE | 3 drug MZ | Labored breathing for 5 minutes; snoring loudly; taking a giant breath; making crying sounds; having stomach movements | 11 h 33 min |
| 9 | FL | 985 g; bloody froth expressed on slight manual compression | 825 g | None noted | 3 drug MZ | Eye and head movements | 16 h 24 min |
| 10 | FL | 750 g; ooze bloody froth with slight manual compression | 595 g | None noted | 3 drug MZ | Visible breathing for 5 minutes | 15 h 24 min |
| 11 | FL | 735 g; acute edema and congestion | 720 g | None noted | 3 drug MZ | Breathing and movement slowed | 13 h 17 min |
| 12 | FL | 815 g; sectioned surfaces ooze blood and frothy fluid | 775 g | None noted | 3 drug MZ | Breathing ceased after 5 minutes | 15 h 14 min |
| 13 | FL | 760 g; pink tinged frothy fluid tracheobronchial tree; frothy fluid oozes from lung surface on compression | 545 g | Acute pulmonary edema and congestion | 3 drug MZ | Left arm and shoulder twitching for 15-30 sec | 14 h 28 min |
| 14 | AR | 835 g; mild diffuse edematous change | 735 g | None noted | 3 drug MZ | Chest moving up and down for at least 5 minutes; cheeks moving for 7 minutes | 1 day |
| 15 | FL | 860 g; slight congestion | 590 g | Mild alveolar edema | 3 drug MZ | None available | 2 days |
| 16 | FL | 840 g; ooze frothy red fluid; acute congestion and edema | 620 g | None noted | 3 drug MZ | Chest heaved and face turned purple | 19 h 49 min |
| 17 | AR | 755 g; white frothy fluid expressed on compression | 660 g | None noted | 3 drug MZ | Appeared to swallow multiple times | 1 day |
| 18 | OK | 740 g; edema | 580 g | Pulmonary edema | 3 drug MZ | Variety of movements including breathing heavily, writhing, mumbling, grimacing, grunting, and straining against restraints | 1 d 13 h 19 min |
| 19 | AL | 430 g; major bronchi contain froth | 600 g | None noted | 3 drug MZ | Breathing quickened; other extremity movements including raising arm | 15 h 12 min |
| 20 | AL | 680 g; mild vascular congestion | 540 g | None noted | 3 drug MZ | Labored breathing for 7 minutes before breathing slowed | 10 h 48 min |
| 21 | AL | 780 g; congested | 980 g | Lungs show mild interstitial anthracosis | 3 drug MZ | Chest moved during early part of execution | 11 h 58 min |
| 22 | VA | 800 g; moderate amount of froth upper airway | 628 g | Lung: within normal limits; scattered hypoxic-ischemic neurons<br>hippocampus | 3 drug MZ | Deep breathing; diaphragm contracted sharply several times | 12 h 15 min |
| 23 | AL | 850 g; congested | 920 g | Pigmented<br>macrophages,<br>mucus plugs<br>in bronchioles | 3 drug<br>MZ | Difficulty breathing, struggling for breath,<br>heaving, asthmatic-sounding barking<br>coughs and gasping respiration; coughed<br>regularly for 15 minutes | 13 h 55 min |
| 24 | OH | 709 g; sections exude<br>serosanguineous frothy<br>fluid | 665 g | Pulmonary<br>edema; HIE | 3 drug<br>MZ | Chest was rapidly rising and falling;<br>labored breathing; puffing lips out as he<br>exhaled; gasping and wheezing loudly | 23 h 46 min |
| 25 | OK | 550 g; edematous; exudes<br>moderate amount of clear<br>frothy fluid | 550 g | Congestion<br>and edema | MZ, PB,<br>KCl | Stated: "My body is on fire" | 16 h 34 min |
| 26 | AR | 590 g; serosanguineous<br>fluid nares, oral cavity,<br>upper and lower airways | 555 g | None noted | 3 drug<br>MZ | Coughing, struggling for air, made noises<br>like gasping and straining for breath;<br>heaving and choking with chest pumping<br>up and down; labored breathing | 1 day |
| 27 | AR | 750 g; abundant watery<br>secretion nares, oral cavity,<br>upper, lower airways | 720 g | None noted | 3 drug<br>MZ | Deep and heavy breathing; sucked in air<br>and repeatedly arched his back over 5-6<br>minutes | 1 day |
| 28 | AZ | 980 g; exude marked<br>amounts of blood and<br>frothy fluid | 945 g | Intra-alveolar<br>hemorrhage<br>and edema<br>fluid<br>bilaterally | MZ +<br>HM | Gasped more than 600 times over 1 hour<br>and 40 minutes; heavy then shallow<br>breathing; struggling efforts to breathe;<br>gulpd "like a fish on land"; snoring and<br>sucking noises | 16 h 14 min |
| 29 | GA | 860 g | 820 g | None noted | PB | N/A | N/A |
| 30 | GA | 720 g; frothy fluid in<br>tracheobronchial tree | 640 g | None noted | PB | N/A | N/A |
| 31 | GA | 860 g; congestion | 720 g | None noted | PB | N/A | N/A |
| 32 | GA | 740 g; moderate<br>congestion | 580 g | None noted | PB | N/A | N/A |
| 33 | GA | 820 g; mild degree frothy<br>fluid lower airways | 720 g | None noted | PB | N/A | N/A |
| 34 | GA | 760 g; cut surface exudes<br>minimal frothy fluid | 700 g | None noted | PB | N/A | N/A |
| 35 | GA | 620 g; moderate amount of<br>white foam upper airway | 520 g | None noted | PB | N/A | N/A |
| 36 | GA | 760 g; congestion and<br>edema | 640 g | None noted | PB | N/A | N/A |
| 37 | GA | 800 g; frothy material main<br>bronchi and small airways | 640 g | None noted | PB | N/A | N/A |
| 38 | GA | 720 g; congestion | 600 g | None noted | PB | N/A | N/A |
| 39 | GA | 580 g; edema | 580 g | None noted | PB | N/A | N/A |
| 40 | GA | 580 g; edema; edematous<br>fluid major airways | 460 g | None noted | PB | N/A | N/A |
| 41 | GA | 520 g; white frothy fluid<br>tracheobronchial tree | 540 g | None noted | PB | N/A | N/A |
| 42 | GA | 760 g | 620 g | None noted | PB | N/A | N/A |
| 43 | GA | 780 g; upper airways<br>moderate amounts of white<br>foam | 680 g | None noted | PB | N/A | N/A |

## Discussion

In this study of autopsy findings of 43 inmates executed by a protocol involving Midazolam or single drug Pentobarbital, we unexpectantly found evidence of lung edema in 34 of 43 cases. Lethal injection is not a medical act and protocol revisions have been driven by drug availability and not as an act of technical refinement or process improvement. Lethal injection has not been subject to the oversight of medical science because of a prohibition of involvement according to medical ethical opinion.^6^

In *Baze v. Rees*, the United States Supreme Court asserts that since capital punishment is constitutional, “it necessarily follows that there must be a means of carrying it out”.^7^ Lethal injection gained public approval based on a perception that death by this method was not cruel. When lethal injection is set next to the electric chair, lethal gas, or the firing squad, it seemed the clear winner as compared to these other methods, witnessing death by lethal injection generally reveals little. Even in that case though, documented “botched” executions reportedly occur at a higher rate than other methods^8^. It is conceivable the courts have also struggled to consider the possibility of an 8^th^ amendment violation in an action that outwardly seemed so bloodless. Although adopted as a non-cruel alternative to other methods of execution, lethal injection has increasingly resulted in problematic executions with serious complications. Though the court seems determined to defend lethal injection as a constitutional method of execution, death row prisoners regard this quite differently. Recent execution jurisprudence has seen a rise in claims by prisoners who now seek to be executed by lethal gas, the firing squad, and the electric chair. In *Bucklew v. Presythe*^9^, Bucklew argued death by lethal gas would be a less cruel method of execution as he suffered from cavernous hemangiomas within his airway. In the majority opinion, the court did not find in favor of this claim. The State of Tennessee employs the three-drug method beginning with Midazolam. Death-row prisoners in Tennessee chose to die by electric chair and not lethal injection as a result of fear about cruelty in the Tennessee lethal injection method (personal communication with Zivot, February 7, 2019). In *Glossip v. Gross*, the Supreme Court referred to 11 executions involving 500 mg of intravenous midazolam that “appear to have been conducted with no significant problems”^10^. Nine of those cases were part of this study, and all showed acute pulmonary edema; there is no autopsy data available for the other two. This statement requires reassessment considering new evidence that adds pulmonary edema to the list of unanticipated complications of lethal injection using midazolam and pentobarbital. The additional likelihood of awareness during execution by the 3-drug midazolam method with attendant sensations of air hunger and drowning raises grave concerns about the suitability of midazolam (a drug which cannot induce general anesthesia) as the first drug in three-drug lethal injection protocols.

According to the 8^th^ amendment of the US constitution, punishment must not be cruel. The evolving definition of cruelty tracks with the evolution of civil society. Witnesses to executions are necessary in order to engage their empathy to judge the cruelty of the act but lethal injection execution witnesses see a state-curated event. According to Austin Sarat^11^, in a review of 1,054 lethal injection executions, witnesses report that the execution appeared botched in 75 cases (7.12%). Before now, physical evidence for cruelty has been lacking and these autopsy findings suggest that the rate of cruelty in lethal injection executions may be significantly higher than what can be seen by traditional visual observation alone.

The autopsy has existed in some form for centuries, and the discipline of pathology as a medical subspecialty has its roots in the modern autopsy as practiced in Central Europe during the 19^th^ century. Autopsy is from the Greek *autopsia* and translates to “see for oneself”.^12^ The alternate term necropsy, means “to look at the dead”.^13^ The purpose of the autopsy has varied considerably over time, but most autopsies performed today are carried out to pinpoint a cause (and sometimes manner) of death and to thoroughly document the causes and effects of disease on the body. An autopsy may be considered the only complete physical examination ever made of a body, and just as antemortem physical examinations vary in thoroughness and specific focus depending on the indication for the procedure, so too do autopsies differ in the level of detail in dissection and description depending on the context in which they are performed. Not all states require autopsies following execution, but some jurisdictions perform autopsies for all homicides. Other purposes for autopsy in this setting include documentation of lethal injection, confirmation of correct intravenous catheter placement, exclusion of any other contributing cause of death, and to search for unexpected complications of the procedure. Autopsies on executed prisoners also have focused on determination of the presence and quantity of execution drugs in post-mortem blood samples. Lethality claims about death by injected pentobarbital drawn from literature suggesting fatality with a blood concentration of 30 mg/L.^14^ Rapid and accurate methods to detect the presence of barbiturates have been available for a number of years but uncertainty remains about what blood level of a barbiturate is necessary to produce death.^15^ Midazolam is a benzodiazepine with low associated toxicity and death by the exclusive use of midazolam is, to our knowledge, unreported.

Our intent in this study was to review a set of autopsy reports (written by pathologists from seven different states) following lethal injection to further our understanding of the process by which death is brought about in this situation. Significant pathology in the lungs emerged as a consistent and unexpected finding. Pulmonary edema and congestion are common findings in hospital autopsies where death is usually a gradual process, often as a result of progressive cardiac failure or overwhelming infection, but it is not seen in cases of sudden death^16^. Likewise, froth in the airways (while commonly seen in acute cardiogenic pulmonary edema) suggests physical or chemical injury as seen in death by drowning, electrocution, inhalation of toxic gas, or heroin overdose. It requires breathing to create froth, so the timing of pulmonary edema must, but definition, occur prior to the administration of a paralyzing agent or heart stopping medication (after which breathing does not occur). The cause of pulmonary edema in heroin overdose has been debated with proposed etiologies including central respiratory depression, hypoxia-induced increase in capillary permeability, primary toxic effect on alveolar capillaries, and myocardial depression^17^.

The finding of “vascular congestion or parenchymal edema” is an alarming one for the question of cruelty in this method of execution. The subjective experience of acute pulmonary edema has been listed for patients partly in these words: “A feeling of suffocating or drowning that worsens when lying down”^18^

If it is assumed that agents used in the 3-drug midazolam protocol function as they do in clinical settings, then midazolam would cause sedation, the paralytic would quickly induce apnea, and potassium chloride would quickly stop the heart. In this sequence of events there is no reason to expect pulmonary congestion or edema; in fact, lung weights close to those seen in very sudden death (i.e. normal weights) would be anticipated. Normal adult lung weights vary according to gender with the right lung generally weighing more than the left. In one paper, the average lung weight was reported as 234 grams (s +/- 39)^19^. In another paper describing the weight of lungs in men, authors report the weight of the right lung to range between 155-720 grams and the left lung to range between 112-675 grams^20^. The lung weights observed in this study were all above 400 grams (average weights for right and left lungs, respectively, in midazolam executions were 787 grams and 707 grams and for pentobarbital executions 723 grams and 631 grams), indicating some combination of vascular congestion and parenchymal edema.

As pentobarbital is not administered to patients in a clinical setting in the massive doses used for execution (e.g. 5 grams), its effects on the body at high concentration are unknown. Pentobarbital is a highly alkaline solution (pH between 9.8-11)^21^. Pentobarbital is a Barbiturate and these drugs have long been known to cause vascular injury if improperly administered, with inadvertent intra-arterial injection sometimes causing gangrene^22^. In contrast, midazolam preparations for injection are adjusted to a more acidic pH of 3 to 3.6 ^23^. It is possible that the massive quantities of acidic or basic solution entering the bloodstream during lethal injection may be directly toxic to pulmonary capillary endothelial cells and that the earliest manifestation of this injury is the escape of edema fluid into the normally air-filled lungs. The statement by inmate 25, “My body is on fire” raises the possibility that intravascular acid-sensing ion channel activation by the large quantity of highly acidic midazolam. Pain at the injection site is reported in cases when the injected solution has a pH less than 4 or greater than 11^24^.

It is intuitive that pulmonary edema in the three-drug midazolam protocol is caused by midazolam itself for several reasons. First, witnesses report respiratory distress early in the procedure (sometimes almost immediately after announcement that the execution has begun), before the paralytic agent or potassium chloride could have taken effect. Second, the production of froth in the setting of pulmonary edema requires the active mixing of fluid, proteins, and surfactant, and this process ceases after the paralytic takes effect; non-aerated edematous lung tissue does not produce froth^25^. Third, intravenous acid is known to cause diffuse lung injury in animal models^26^, a process that begins with exudation of fluid and protein into the alveolar space. Pulmonary edema in this setting is appropriately described as “flash pulmonary edema” in contrast with that accompanying death from natural causes (agonal pulmonary edema). Furthermore, even in massive overdoses, benzodiazepines very rarely cause death and would not be expected to cause pulmonary edema as an agonal affect associated with gradual multi-organ and cardiac failure.

We speculate that pentobarbital produces pulmonary edema via a similar mechanism, specifically the caustic action of alkaline solution on pulmonary alveolar capillaries, but because of the different mechanism of action of pentobarbital which includes a direct respiratory depressant effect, it is difficult to be confident of this. As indicated above, heavy, congested, edematous lungs are common following death from a wide variety of causes, but fulminant pulmonary edema with froth filling the respiratory tree is a noteworthy finding more commonly associated with lung injury than pure respiratory depression.

There were a number of limitations inherent in this study. First, the authors did not “see for themselves” firsthand the anatomic findings detailed above (except in case 24), but rather relied on written autopsy reports. This has the advantage of limiting biased observations as pathologists from eight different states recorded consistent findings of pulmonary edema, but also makes it possible that minor degrees of pulmonary edema may have been overlooked if not specifically sought. Similarly, histologic examination (which would have afforded a second opportunity to detect mild pulmonary edema) was performed in a minority of cases. It is also true that detailed medical histories seeking conditions which might predispose to pulmonary edema were not collected on these inmates, but it would be extraordinary if the majority of subjects in this series suffered from a medical predisposition to acute pulmonary edema.

Negative pressure pulmonary edema (caused by the development of a very high negative intrathoracic pressure) can develop as the result of airway obstruction in the context of sedation, but this typically manifests as a struggle for breath against upper airway obstruction followed by a deep inspiration. In the setting of negative pressure pulmonary edema, the observed lung fluid has a low protein concentration suggesting a hydrostatic, as opposed to a direct toxic effect.^27^ It is not clear if the pulmonary edema fluid observed in our review had high or low levels of protein. Witnesses to executions, including one of the authors (JZ), do not report this pattern of breathing, making it unlikely to have played a major role in these cases.

## Conclusion

In our review of the autopsy findings of 43 inmates executed by a protocol involving Midazolam or single drug Pentobarbital, we found evidence of lung edema in 34 of 43 cases (79%) The occurrence of pulmonary edema in the setting of lethal injection execution may not be easily observed by witnesses to execution. Pulmonary edema is an extremely uncomfortable experience^28^, and these findings raise the concern that lethal injection execution is far more uncomfortable an experience than previously believed. The mechanism of the development of pulmonary edema in these case remains uncertain, but it is possibly the result of the injection of large quantities of unphysiological pH solutions. The protein content in the observed pulmonary edema fluid was not measured and further analysis of this fluid would be warranted in order to determine if the mechanism of pulmonary edema was hydrostatic as opposed to cytotoxic.

## Data Availability

All data produced in the present study are available upon reasonable request to the authors.

